# Immunohistochemical phenotype is associated with metastatic site in breast cancer: a retrospective pathomorphological study of women from the Lower Aral Sea region, Uzbekistan

**DOI:** 10.64898/2026.06.05.26354969

**Authors:** Adham A. Khodjaniyazov, Ruslon R. Rojobov

## Abstract

**Background:** Breast cancer is the most frequently diagnosed cancer and the leading cause of cancer death in women worldwide, and the great majority of these deaths are caused by metastatic disease. Whether the immunohistochemical (IHC) phenotype of breast cancer is associated with the anatomical site of metastasis has been characterized mainly in high-income, registry-based populations, while data from ecologically stressed and medically under-served regions such as the Lower Aral Sea basin are lacking.

**Methods:** We retrospectively reviewed 652 women diagnosed with breast cancer at the Khorezm Branch of the Republican Specialized Scientific-Practical Medical Center of Oncology and Radiology (Uzbekistan) between 2020 and 2024, of whom 213 had metastatic disease (306 metastatic foci). Histological type was assessed on hematoxylin–eosin and van Gieson–stained sections; quantitative morphometry was performed in Fiji/ImageJ; and HER2, estrogen receptor (ER), progesterone receptor (PR) and Ki-67 were assessed by IHC. The association between marker expression and metastatic site (liver, lung, lymph node) was tested in 187 foci with adequate tissue using the chi-square test, with significance at p < 0.05.

**Results:** Invasive ductal carcinoma predominated. Metastatic site was significantly associated with the IHC phenotype. Liver metastases showed the highest frequency of HER2 3+ (45.7%), ER-negativity (65.2%), PR-negativity (69.6%) and high proliferation (Ki-67 ≥ 60%; 47.8%), whereas lymph-node metastases were more often hormone-receptor-positive (ER+ 58.7%; PR+ 52.4%) with lower HER2 3+ (22.2%); lung metastases were intermediate (all p < 0.05). The combination of HER2 3+ and Ki-67 ≥ 60% was associated with multi-organ spread. Morphometry corroborated these patterns: liver lesions had larger atypical cells (up to 132.8 μm), a higher nuclear-to-cytoplasmic ratio (0.76 vs 0.51) and more extensive necrosis and microvascularity than lymph-node lesions. A pragmatic 5-criterion morphological score (histological type, Ki-67, HER2, ER/PR status, atypical-cell size) stratified metastatic risk into three tiers.

**Conclusions:** In this regional cohort, the IHC phenotype of breast cancer tracked the anatomical site of metastasis, with an aggressive HER2-driven, hormone-receptor-negative profile concentrated in liver metastases and a hormone-receptor-positive profile in lymph-node metastases. These findings reproduce established organotropism patterns in a previously uncharacterized population and support phenotype-aware, site-specific surveillance together with a low-cost morphological risk score for resource-limited settings.

## 1. Introduction

Breast cancer is the most frequently diagnosed malignancy and the leading cause of cancer death among women worldwide; in 2022 an estimated 2.3 million new cases were recorded globally, and the disease ranks first for both incidence and mortality in women in the great majority of countries [1]. Despite advances in early detection and systemic therapy, a substantial proportion of patients present with, or progress to, advanced disease, and metastasis accounts for the overwhelming majority of breast-cancer deaths [2,3]. In the population from which the present cohort was drawn, the incidence of breast cancer has been rising by approximately 1.8% per year, with an estimated cumulative risk of about 5.4% by 74 years of age.

Breast cancer is biologically heterogeneous. Routine immunohistochemical (IHC) assessment of the estrogen receptor (ER), the progesterone receptor (PR), human epidermal growth factor receptor 2 (HER2) and the proliferation marker Ki-67 defines surrogate molecular phenotypes that guide prognosis and treatment selection and that also shape metastatic behavior [2,6]. The same panel underpins decisions about endocrine therapy, HER2-directed agents and cytotoxic chemotherapy in routine practice.

Metastatic dissemination is not random. Breast cancer cells preferentially colonize particular organs — a phenomenon termed organotropism — that is governed by tumor-intrinsic molecular features and by the receptivity of the distant-organ microenvironment [3]. Across large registry and clinical series, hormone-receptor-positive tumors tend to spread to bone, HER2-enriched tumors show a liver-(and brain-) homing tendency, and triple-negative tumors favor the lung [3,4,5]. Recognizing these associations enables phenotype-aware surveillance and individualized management of patients at risk of visceral relapse.

Most of this evidence, however, derives from registry data in high-income settings. Populations living in ecologically stressed, medically under-served regions remain poorly characterized at the morphological level. The Lower Aral Sea basin — comprising the Republic of Karakalpakstan and the Khorezm region of Uzbekistan — has been affected by one of the most severe man-made ecological disasters of the modern era, yet the morphological and immunophenotypic features of metastatic breast cancer in women of this region have not been systematically described [10,11].

We therefore conducted a retrospective pathomorphological study with two aims: first, to characterize the histological and quantitative morphometric features of metastatic breast cancer in women of the Lower Aral Sea region; and second, to test whether the IHC phenotype of the tumor is associated with the anatomical site of metastasis.

## 2. Materials and methods

### 2.1 Study design and population

This was a single-center, retrospective study based on the clinical and pathology archives of the Khorezm Branch of the Republican Specialized Scientific-Practical Medical Center of Oncology and Radiology. All women registered with a diagnosis of breast cancer between 2020 and 2024 were eligible. A total of 652 cases of breast cancer were reviewed, of which 213 (32.7%) had documented metastatic disease. Among these 213 women, 306 separate metastatic foci were identified, indicating a predominance of multi-organ involvement. Metastases were most frequent in the 45–59-year age group. Within the region, the highest case numbers came from the city of Urgench (n = 37), Urgench district (n = 27), Shovot district (n = 20) and Khonqa district (n = 20), which together accounted for roughly 40% of cases. Surgical treatment had been performed in 156 of the 213 women (73.2%); in 57 (26.8%) surgery had not been undertaken for various clinical reasons.

### 2.2 Histopathology and morphometry

Diagnostic material comprised biopsy specimens and archival paraffin blocks. Sections of 5– 7 μm were stained with hematoxylin–eosin for routine assessment and with van Gieson stain for evaluation of connective tissue. Tumors were classified by histological type as invasive ductal carcinoma, invasive lobular carcinoma or mixed forms. Quantitative morphometry was performed in Fiji (ImageJ). Measured parameters included atypical-cell size, the nuclear-to-cytoplasmic ratio, fibrosis coverage, total collagen-fiber area, necrosis-focus area, the number of microvessels per high-power field and the epithelium-to-stroma ratio. Because adequate metastatic tissue could not be retrieved for every site, detailed morphometric measurement was confined to liver (n = 6) and lymph-node (n = 7) biopsy material; lung metastases were evaluated within the immunohistochemical analysis.

### 2.3 Immunohistochemistry

Immunohistochemistry was performed for HER2/neu, ER, PR and Ki-67 using standard protocols with reagents from Dako and Leica. Antigen–antibody reactivity was graded on a 0 to 3+ scale; HER2 was scored according to the principles of the ASCO/CAP guideline [7], ER and PR according to the ASCO/CAP recommendations for hormone-receptor testing [8], and Ki-67 in line with the recommendations of the International Ki-67 in Breast Cancer working group [9]. Across the metastatic cohort, strong HER2 expression (3+) was present in 58 patients (27.2%), moderate (2+) in 32 (15.0%), weak (1+) in 41 (19.2%) and negative in 82 (38.5%). ER positivity was recorded in 117 patients (54.9%) and PR positivity in 104 (48.8%). A high proliferative index (Ki-67 > 20%) was seen in 143 patients (67.1%).

### 2.4 Metastatic-risk score

To integrate morphological and immunophenotypic information, a 5-criterion scoring scheme was constructed combining histological type, Ki-67 index, HER2 expression, hormone-receptor status and atypical-cell size, with each criterion scored from 1 to 3 points (Table 3). Total scores range from 5 to 15 and define three risk tiers.

### 2.5 Statistical analysis

Data were analyzed in SPSS 26.0 and STATISTICA for Windows. Associations were examined using correlation analysis and logistic regression. Differences between groups were assessed with the Student t-test, the chi-square (χ^2^) test and the Fisher exact test. A two-sided p-value below 0.05 was considered statistically significant.

### 2.6 Ethics

The study used anonymized archival diagnostic material. Ethical approval was obtained from the relevant institutional ethics committee ethics committee of the Urgench State Medical Institute (approval No. 87 dated 12.04.2024); because the analysis was based on de-identified archival specimens, the requirement for individual informed consent was waived in accordance with local regulations.

## 3. Results

### 3.1 Cohort characteristics and histological type

Invasive ductal carcinoma accounted for the largest share of metastatic breast cancers in the cohort. These tumors retained only partial glandular architecture and showed epithelial atypia, vacuolated cytoplasm and hyperchromatic nuclei, with lymphoid infiltrates and areas of fibrosis in the stroma and infiltration of adjacent adipose tissue. Invasive lobular carcinomas had largely lost glandular architecture, with cells arranged in single-file (chain-like) patterns; nuclei were relatively uniform in size and shape but cytoplasm was scant, and cell–cell adhesion was reduced. Mixed tumors displayed a heterogeneous appearance combining ductal and lobular features, with stromal sclerosis predominating and only weakly formed invasive ductal changes.

### 3.2 Morphometry of liver and lymph-node metastases

Morphometric analysis revealed a clear quantitative distinction between liver and lymph-node metastases (Table 1). Liver lesions contained the largest atypical cells (up to 132.8 μm), the highest nuclear-to-cytoplasmic ratio (0.76 ± 0.11), the most extensive necrosis (4720 ± 1380 μm^2^) and fibrosis (30.8 ± 6.4%), and the greatest microvessel density (12.2 ± 3.4 per high-power field), with hyperchromatic nuclei and reactive fibrosis as characteristic features. Lymph-node lesions comprised smaller cells (up to 68.59 μm), a lower nuclear-to-cytoplasmic ratio (0.51 ± 0.08) and less necrosis (2180 ± 740 μm^2^), with sinus obstruction and hyalinosis predominating. Together, these measurements indicate a structurally more aggressive profile in liver metastases.

**Table 1.** Morphometric indicators of liver and lymph-node metastases. Values are the range, the median, or the mean ± standard deviation. Measurements were obtained in Fiji/ImageJ; lung metastases were not measured because of insufficient biopsy material.

| Morphometric indicator | Liver (n = 6) | Lymph node (n = 7) |
| --- | --- | --- |
| Atypical-cell size range ($\mu\text{m}$) | 20.0–132.8 | 19.85–68.59 |
| Median cell size ($\mu\text{m}$) | 64.5 | 33.8 |
| Nuclear-to-cytoplasmic ratio | $0.76 \pm 0.11$ | $0.51 \pm 0.08$ |
| Fibrosis coverage (%) | $30.8 \pm 6.4$ | $21.6 \pm 5.2$ |
| Collagen-fiber area (%) | $18.6 \pm 5.1$ | $14.2 \pm 4.3$ |
| Necrosis-focus area ($\mu\text{m}^2$) | $4720 \pm 1380$ | $2180 \pm 740$ |
| Microvessels per high-power field | $12.2 \pm 3.4$ | $6.2 \pm 2.1$ |
| Epithelium-to-stroma ratio | $1.78 \pm 0.38$ | $1.10 \pm 0.24$ |
| Characteristic morphology | Hyperchromatic nuclei;<br>reactive fibrosis | Sinus obstruction;<br>hyalinosis |

### 3.3 Immunohistochemical marker expression

Marker expression varied with histological type. Strong HER2 (3+) expression was significantly more frequent in invasive ductal carcinomas than in lobular carcinomas (p < 0.05) and was accompanied by aggressive morphology, marked disturbance of the nuclear-to-cytoplasmic ratio, reduced intercellular adhesion and high mitotic activity. Lobular carcinomas more often showed ER and PR positivity, with strong nuclear staining confirming hormone dependence. Mixed tumors had a heterogeneous immunophenotype with features of both components. The Ki-67 index separated tumors by biological aggressiveness: tumors with Ki-67 ≥ 60% (68 patients; 31.9%) showed rapid growth and a high metastatic propensity, those with Ki-67 of 30–50% (75 patients; 35.2%) followed a more indolent course, and those with Ki-67 < 20% (70 patients; 32.9%) carried a comparatively favorable prognosis. ER-negative/PR-negative tumors frequently co-expressed HER2 and high Ki-67 and were characterized by aggressive behavior.

### 3.4 Association of immunophenotype with metastatic site

The relationship between marker expression and metastatic site was tested in the 187 foci with adequate tissue (liver, n = 46; lung, n = 78; lymph node, n = 63). All four markers were significantly associated with metastatic site (Table 2). Liver metastases showed the highest frequency of HER2 3+ (45.7%) and of high proliferation (Ki-67 ≥ 60%; 47.8%) together with the greatest hormone-receptor negativity (ER-negative 65.2%; PR-negative 69.6%), defining an aggressive, HER2-driven, hormone-receptor-negative profile. Lymph-node metastases were more often hormone-receptor-positive (ER+ 58.7%; PR+ 52.4%) with lower HER2 3+ (22.2%), indicating an association with hormone-dependent tumors. Lung metastases occupied an intermediate position (HER2 3+ 28.2%; ER+ 48.7%; Ki-67 ≥ 60% 35.9%). Overall, the combination of HER2 3+ and Ki-67 ≥ 60% was associated with a higher frequency of multi-organ metastasis (p < 0.05), whereas ER-positive/PR-positive tumors followed a comparatively slower metastatic course.

**Table 2.** Association between immunohistochemical marker expression and metastatic site (n = 187 foci). The χ^2^ and p values apply to each marker. Foci in bone, brain and other sites (119 of 306) were excluded because biopsy material was insufficient.

| Marker / status | Lung (n = 78) | Lymph node (n = 63) | Liver (n = 46) | $\chi^2$ | p |
| --- | --- | --- | --- | --- | --- |
| HER2 3+ | 22 (28.2%) | 14 (22.2%) | 21 (45.7%) | 7.82 | < 0.05 |
| HER2 0 / 1+ / 2+ | 56 (71.8%) | 49 (77.8%) | 25 (54.3%) |  |  |
| ER positive | 38 (48.7%) | 37 (58.7%) | 16 (34.8%) | 6.94 | < 0.05 |
| ER negative | 40 (51.3%) | 26 (41.3%) | 30 (65.2%) |  |  |
| PR positive | 34 (43.6%) | 33 (52.4%) | 14 (30.4%) | 5.68 | < 0.05 |
| PR negative | 44 (56.4%) | 30 (47.6%) | 32 (69.6%) |  |  |
| Ki-67 ≥ 60% | 28 (35.9%) | 18 (28.6%) | 22 (47.8%) | 4.52 | < 0.05 |
| Ki-67 30–50% | 29 (37.2%) | 25 (39.7%) | 14 (30.4%) |  |  |
| Ki-67 < 20% | 21 (26.9%) | 20 (31.7%) | 10 (21.7%) |  |  |

### 3.5 Metastatic-risk score

The 5-criterion score is summarized in Table 3. Combining histological type, Ki-67 index, HER2 expression, hormone-receptor status and atypical-cell size, total scores ranged from 5 to 15 and defined three tiers: high risk (12–15 points), indicating a high probability of rapid metastasis and warranting aggressive treatment; moderate risk (8–11 points), warranting close monitoring and combination therapy; and low risk (5–7 points), associated with a comparatively favorable prognosis and standard treatment protocols.

**Table 3.**
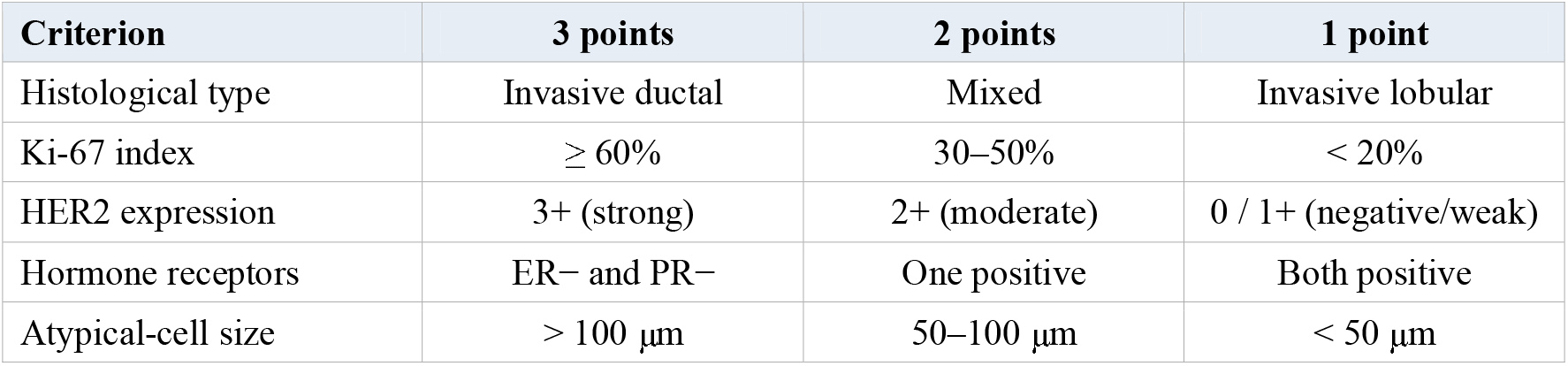
Five-criterion morphological score for stratifying metastatic risk. Each criterion contributes 1–3 points (total 5–15): high risk, 12–15; moderate risk, 8–11; low risk, 5–7.

| Criterion | 3 points | 2 points | 1 point |
| --- | --- | --- | --- |
| Histological type | Invasive ductal | Mixed | Invasive lobular |
| Ki-67 index | $\geq 60\%$ | 30–50% | $< 20\%$ |
| HER2 expression | 3+ (strong) | 2+ (moderate) | 0 / 1+ (negative/weak) |
| Hormone receptors | ER– and PR– | One positive | Both positive |
| Atypical-cell size | $> 100 \mu\text{m}$ | 50–100 $\mu\text{m}$ | $< 50 \mu\text{m}$ |

## 4. Discussion

In this retrospective cohort of women with metastatic breast cancer from the Lower Aral Sea region, the immunohistochemical phenotype of the tumor was significantly associated with the anatomical site of metastasis. Liver metastases concentrated an aggressive profile — frequent HER2 overexpression, hormone-receptor negativity and high proliferation — whereas lymph-node metastases were enriched for hormone-receptor-positive tumors, and lung metastases were intermediate. The morphometric data reinforced this picture, showing larger atypical cells, a higher nuclear-to-cytoplasmic ratio and more extensive necrosis and microvascularity in liver lesions than in lymph-node lesions.

These findings are concordant with the established biology of breast-cancer organotropism. Large registry and clinical analyses have repeatedly shown that HER2-enriched tumors carry a liver-(and brain-) homing tendency, that hormone-receptor-positive tumors are comparatively indolent and bone- or node-associated, and that triple-negative tumors favor the lung [3,4,5]. The aggregation of HER2 3+ with high Ki-67 in liver metastases, and the association of this combination with multi-organ spread, is consistent with the more aggressive natural history of HER2-driven and highly proliferative disease [3,4]. The present study extends this body of evidence by demonstrating that the same phenotype–site relationships are recoverable in a previously uncharacterized, ecologically stressed and medically under-served population, using routinely available histology and a four-marker IHC panel.

The results carry practical implications for resource-limited settings. First, they support phenotype-aware, site-specific surveillance: women whose primary tumors are HER2-positive, hormone-receptor-negative and highly proliferative warrant heightened vigilance for visceral — particularly hepatic — relapse, while hormone-receptor-positive disease may justify a different surveillance emphasis. Second, the pragmatic 5-criterion morphological score offers a low-cost means of stratifying metastatic risk where comprehensive molecular profiling is not available. Third, integration of morphological and immunophenotypic data through telepathology can extend specialist pathology to remote districts of the region, improving the timeliness and consistency of diagnosis.

Several limitations should be acknowledged. The study was retrospective and single-center, with the attendant risks of selection and information bias. Morphometric measurement was restricted to small subsets of liver (n = 6) and lymph-node (n = 7) biopsies, so those quantitative estimates should be interpreted with caution. Because reliable metastatic tissue could not be obtained for every site, the marker-by-site analysis relied predominantly on the primary tumors of patients with documented metastases, and 119 of the 306 foci (in bone, brain and other organs) were excluded for lack of material; the site associations therefore describe the three sampled organs rather than the full metastatic landscape. Subtyping was based on a surrogate IHC panel rather than full molecular profiling, and equivocal HER2 (2+) results were not routinely confirmed by in-situ hybridization. Finally, the study did not include survival follow-up, so the prognostic value of the phenotype–site associations and of the proposed score requires prospective, multi-center validation with clinical outcomes.

## 5. Conclusions

In women with metastatic breast cancer from the Lower Aral Sea region of Uzbekistan, the immunohistochemical phenotype of the tumor was associated with the anatomical site of metastasis: an aggressive HER2-driven, hormone-receptor-negative, highly proliferative profile was concentrated in liver metastases, while a hormone-receptor-positive profile characterized lymph-node metastases. These regional findings reproduce established organotropism patterns in a previously uncharacterized population and provide a rationale for phenotype-aware, site-specific surveillance and for a simple, low-cost morphological risk score suited to resource-limited settings. Prospective studies linking these features to clinical outcomes are warranted.

## Data Availability

All data produced in the present study are available upon reasonable request to the authors.

## Declarations

### Ethics approval and consent to participate

The study used anonymized archival diagnostic material. Ethical approval was obtained from the relevant institutional ethics committee [87]; because the analysis was based on de-identified archival specimens, the requirement for individual informed consent was waived in accordance with local regulations.

### Consent for publication

Not applicable.

### Availability of data and materials

The datasets generated and analyzed during the current study are available from the corresponding author on reasonable request.

### Authors’ contributions

A.A.K. conceived and designed the study, collected and analyzed the histological, morphometric and immunohistochemical data, and drafted the manuscript. R.R.R. contributed to the study design, data interpretation and critical revision of the manuscript. Both authors read and approved the final manuscript.

## Acknowledgements

The authors thank the staff of the Khorezm Branch of the Republican Specialized Scientific-Practical Medical Center of Oncology and Radiology for access to clinical and pathology archives.

